# Uncertainty stress, and its impact on disease fear and prevention behaviors during the COVID-19 epidemic in China: A panel study

**DOI:** 10.1101/2020.06.24.20139626

**Authors:** Xiaozhao Yousef Yang, Sihui Peng, Tingzhong Yang, Weifang Zhang, Huihui Wang, Randall R Cottrell

## Abstract

**Objective:** To examine changing trends of uncertainty stress, and its impact on disease fear and prevention behaviors during the Chinese COVID-19 epidemic using a prospective observational study.

**Methods:** The study employed a longitudinal design. Participants were recruited for an online panel survey from chat groups on social media platforms. There were 5 waves of interviews. Information on uncertainty stress and related variables were collected via the online survey. Descriptive statistics and the GIM program were used for data analysis.

**Results:** Participants numbered 150 for the linkable baseline survey and 102 (68%) for the final survey. Uncertainty stress(β: -0.047, S.E: 0.118, p>0.05) did not show a statistically significant temporal change trend over the observation period. Disease fear manifested a statistically significant downwards trend (β: -0.342, S.E: 0.157, p<0.05), and prevention behaviors indicated an upwards trend (β: 0.048, S.E: 0.021, p<0.05) during the observation period. Uncertainty stress was positively associated with disease fear (β: 0.45046, S.E: 0.05964, p<0.0001), and negatively associated with self-efficacy (β: -0.6698, S.E: 0.01035, p<0.0001), and prevention behaviors (β:-0.02029, S.E: 0.00876, p: 0.0209).

**Conclusion:** This study yielded new information about uncertainty stress among Chinese people during the COVID-19 epidemic. Policy changes and public education are essential for minimizing the negative effects of uncertainty stress in disease prevention.

## Introduction

The COVID-19 pandemic represents a massive global health crisis (World Health Organization, 2020). Among the characteristic of this outbreak are the unpredictability of the situation, the uncertainty of when the disease may be controlled and the seriousness of the risk. These, along with misinformation about the pandemic outbreak, mistrust in the public health system, and the underprepared health facilities to address COVID-19 caused great mental stress (Petterson et al, 2020; Zandifar & Badrfam, 2020; Greenberg et al, 2020). This is a new novel virus and much is unknown. These are unprecedented times, and uncertainty may lead to fear which may give way to dread (Petterson, 2020).

Uncertainty is one of the predominant psychological state that occurs among individuals who are affected directly or influenced by the mixed information about an epidemic. Uncertainty occurs when an event or a situation causes ambiguity, inconsistency, or unpredictability (Mast, 1995). Unfortunately, on top of the ambiguous and idiosyncratic feelings of psychological uncertainty, uncertainty also constitute a stressor in people’s life. A host of evidence supports the assertion that uncertainty constitutes a powerful stressor (Dar et al, 2017; Greco & Roger, 2003; Yang, 2018). Some studies have found that uncertainty stress has a more harmful impact on health than normal life stress (Yang et al, 2018; Wu et al, 2020). Festinger hypothesized that uncertainty causes cognitive confusion (Festinger 1957). When people are confronted with uncertainty stress, they tend to engage in avoidance behavior instead of seeking adaptive measures to tackle recognizable problems. When uncertainty stressors cannot be managed well, mental and behavioral problems may occur over time. Uncertainty stress associated with the COVID-19 epidemic may affect the public’s willingness to learn about and adopt and prevention behaviors to reduce the risk of COVID-19 infection. Some authors have noted that uncertainty associated with the COVID-19 epidemic has created challenges for disease prevention (Petterson et al, 2020; Brown, 2020).

Beyond uncertainty, self-efficacy may form a particular dimension, correlated to but distinct from uncertainty, to affect the effectiveness of adopting prevention behaviors. The Theory of Planed Behavior posits that self-efficacy is a key element in behavioral adaptation (Yang, 2018). Thus self-efficacy for preventing COVID-19 may have an important influence on mental and behavioral responses during the epidemic. Some studies on acute respiratory infections indicate that self-efficacy has an important role in prevention (Ingram et al, 2013 Cheng & Wong, 2005, Perrin et al, 2009).

However, there is currently no empirical research on uncertainty stress from the COVID-19 epidemic.

This study will examine uncertainty stress issues from COVID-19 epidemic. There are three key objectives to this study:

1. To examine levels of uncertainty stress among Chinese citizens amidst the COVID-19 epidemic.
2. To evaluate temporal changing trends in uncertainty stress and related variables during the epidemic.
3. To study the associations between uncertainty stress, disease fear, prevention behaviors, and self-efficacy for preventing the disease.

This study may yield important information for formulating policy and public education initiatives aimed at reducing the disease infection rate, while targeting effective interventions for preventing and mitigating COVID-19.

## Methods

### Study design

A prospective longitudinal panel study was designed to examine temporal trends and changes in uncertainty stress, disease fear, and prevention behavior during the COVID-19 pandemic.

Participants were recruited from social media groups on WeChat and Douban. Several inclusion criteria were utilized in this study. A special administrative WeChat group was established to manage the follow-up data collection, using a unique QR code for each respondent. After scanning the QR code, survey participants could enter the investigation group without further preconditions. This panel study analyzed five waves of data collected: wave 1(5/Feb/2020), wave 2(12/Feb/2020), wave 3(19/Feb/2020), wave 4 (26/Feb/2020), and wave 5(4/March/2020). The entire observation period covered the peak and trough of the COVID-19 epidemic in China. Confirmed new patients respectively numbered 3,887, 2,015, 394, 433, and 133 at the time of each wave (National Health Commission of People’s Republic of China, 2020). The details see reference (Yang et al, 2020).

### Data Collection

An online survey was implemented on an Internet survey platform *Wenjuanxing* (www.wjx.cn). Each wave of the survey had a dedicated electronic questionnaire access link. Repeated questionnaires were distributed during 9:00-11:00 am every Monday. The details see reference (Yang et al, 2020). This study was approved by the ethics committee of Zhejiang University.

### Measurement

In this study, basic individual demographic characteristics were recorded as age, gender, ethnicity, education level, marital status and occupation.

Uncertainty stress was measured and revised from a previous scale designed by Yang and colleagues (Yang et al, 2019), which have demonstrated acceptable validity, and have since been used extensively in Chinese research (Yang et al 2019; Wu et al. 2020). This instrument has also demonstrated acceptable reliability with a Cronbach’s coefficient alphas of 0.89. It covered four items on uncertainty in various arenas of social and personal life, including current uncertainty about life, social change uncertainty, goal-attainment uncertainty and uncertainty about values. Respondents rated these items on a five-point scale from feel no stress (0), a little stress (1), some stress (2), considerable stress (3), and very strong stress (4). A total stress score for the life stress and uncertainty questionnaire was obtained by adding up the responses to the individual questions. The higher the total score, the greater the perceived level of stress. Consistent with prior practice, a cut off score of 12 or more stress was classified as a higher score and indicated severe uncertainty stress (Yang et al. 2018;Wu et al. 2016).

Several related variables were measured as part of this study. Disease fear refers to people’s “worry level” for the COVID-19 infection, which came from two key concepts in the Health Belief Model, perceptions of risk and severity of risk (Yang, 2018). Perceived risk was measured by a question “I feel that I am always at risk of infection?” Responses were on a 5-point Likert-type from “strongly disagree” to “strongly agree”. Perceived severity was measured by the question, “If you were infected with COVID-19, it would be a serious misfortune.” Responses were again on a 5-point Likert-type scale ranging from “strongly disagree” to “strongly agree”. The results of the above two variables are multiplied to form the disease fear. Prevention behavior against COVID-19 infection was measured by a question, “I feel that I can prevent a COVID-19 infection”, with options on a 5-point Likert-type ranging from “strongly disagree” to “strongly agree”. Self-efficacy for preventing COVID-19 infection was measured by a question “I feel I can avoid infection through prevention”, Responses were on a 5-point Likert-type from “no confidence at all” to “very confident”.

### Data analysis

All data were entered into a database using Microsoft Excel. They were then imported into SAS (9.3version) for statistical analysis. Across survey waves, mean scores were calculated for uncertainty stress and related variables at different observation points. The GIM program was used to conduct repeated measures analysis of variance to determine changing trends across the five observation points, and to examine the association between uncertainty stress and the disease fear, self efficacy, and the prevention behavior using the Armitage linear test (SAS Institute Inc, 2011).

## Results

One hundred and fifty participants were recruited at baseline. The baseline was linkable and there were three intermediate and a final observation point, with 102 (68%) participants remaining for all repeated waves. The respondents were located in 24 provinces across China. Of 102 participants, 61.8% were female and 93.3 % were Han Chinese. The average age of participants was 39.1 years (SD: 12.5), 43.0% were never married, 50.0% were married, others were divorce or widowed. 49.0% of them were high school or junior college graduates, and 51.0% completed undergraduate school or higher. As to occupation, 25.5% were managers, 39.2% were professionals, and 27.5% had other types of occupations.

Table 1 indicated that uncertainty stress did not change significantly over the observation period (β: -0.047, S.E: 0.118, p>0.05). This was also true for self-efficacy (β: 0.014, S.E: 0.025, p>0.05). There was a statistically significant downwards trend in the disease fear (β: -0.342, S.E: 0.157, p<0.05) and an upward trend in prevention behaviors (β: 0.048, S.E: 0.021, p<0.05).

**Table 1.** Means of the variables

| Group | N | Uncertainty stress | Disease fear | Self-efficacy | Prevent behavior |
| --- | --- | --- | --- | --- | --- |
| Time1 | 102 | 9.45(3.58) | 8.56(5.83) | 4.01(0.97) | 3.22(0.77) |
| Time2 | 102 | 9.97(3.67) | 7.54(4.78) | 4.00(0.97) | 3.19(0.63) |
| Time3 | 102 | 9.84(3.66) | 7.52(4.89) | 3.99(0.84) | 3.27(0.71) |
| Time4 | 102 | 9.67(3.84) | 7.27(4.59) | 4.06(0.76) | 3.34(0.59) |
| Time5 | 102 | 9.37(3.88) | 6.98(4.10) | 4.05(0.81) | 3.40(0.71) |
| Mean(SD) | 510 | 9.66(3.76) | 7.58(4.88) | 4.03(0.84) | 3.28(0.68) |
| Trend test($\beta$<S.E) | | -0.047(0.118) | -0.342(0.157)* | 0.014(0.025) | 0.048(0.021)* |

\* $p < 0.05$ , \*\* $p < 0.01$

Table 2 shows that uncertainty stress was positively associated with disease fear(β: 0.45046, S.E: 0.05964, p<0.0001), and negatively associated with self-efficacy (β:-0.6698, S.E: 0.01035, p<0.0001), and prevention behaviors (β: -0.02029, S.E: 0.00876, p: 0.0209).

**Table 2.** Uncertainty stress influencing on disease fear, self-efficacy, and prevent behavior

| Group | $\beta$ | S.E | T Value | Pr > t |
| --- | --- | --- | --- | --- |
| Uncertainty stress->disease fear | 0.45046 | 0.05964 | 7.55 | <0.0001 |
| Uncertainty stress-> self-efficacy | -0.6698 | 0.01035 | -0.645 | <0.0001 |
| Uncertainty stress->prevent behavior | -0.02029 | 0.00876 | -2.32 | 0.0209 |

## Discussion

Addressing a gap in the literature, this study found no significant temporal change in uncertainty stress during the study time period. The average uncertainty stress score was 9.45 at base line, and 9.66 at the last observation point, the highest score was 9.84 at the middle observation point. The uncertainty stress score was fairly consistent over the course of the study and did not decrease even though the epidemic was declining. This indicates people’s general uncertainty stress regarding their life’s direction and the social system were unrelated to the number of new confirmed patients. The average value (9.66<95%C.I: 9.54, 9.78>) of uncertainty stress during the observation period was significantly higher than that (8.16<95% C.I: 8.05, 8.27>) of community residents at ordinary times (Yang et al, 2007). Moreover, one fifth of the people in the study were in a state of high uncertainty stress throughout the observation period. This indicates that uncertainty stress levels from the COVID-19 epidemic impacted large numbers of people and could have significant implications on their health. Studies have found that uncertainty constitutes a powerful stressor (Greco and Roger 2003, Yang, 2018), and may lead to mental problems (Yang et al, 2018; Wu et al, 2020). Uncertainty causes cognitive confusion (Festinger 1957), and has a negative influence on people’s behavior. Uncertainty stresses during the COVID-19 pandemic could pose a major challenge to people’s mental and behavioral health, rendering the mission of organizating and coordinating the reopen of the economy even more challenging. Self efficacy is commonly defined as the belief in one’s capabilities to achieve a goal or an outcome. This contruct also remained relatively stable during the study period.

While uncertainty stress levels remained relatively stable, fear of the disease declined over the observation period. This trend mirrored the decline in new confirmed patients. As number of patients decreased fear decreased. According to the Stimulus, Cognition and Response (SCR) theory, when COVID-19 started to spread (Stimulus) and people became aware of the serious threat the disease posed (Cognition), fear levels were increased (Response). This is consistent with findings from other studies (Li et al., 2020; Lunn et al., 2020; LV et al., 2008; Cullen, 2020, Mak et al, 2009). As the number of cases dropped, the stimulus was reduced and fear was also reduced. Preventive behavior, however, increased significantly over the period of the study. This trend is in the opposite direction of the epidemic. This may be explained as preventive behavior is initiated after people are stimulated, and then gradually strengthens (Yang, 2018). The increase in preventive behavior may also be partially responsible for the reduction in fear.

This study found uncertainty stress was positively associated with disease fear. At the same time new confirmed patients was also positively associated with disease fear(β:-0.00035111 S.E: 0.00014206, p<0.05). But new confirmed patients were not associated with uncertainty stress. This may indicate that new confirmed patients and uncertainty stress, are two independent factors in relation to disease fear.

The study also found that uncertainty stress was negatively associated with both self-efficacy and preventive behavior. A host of evidence now supports the assertion that uncertainty is even more stressful than knowing the inevitability of something bad happening (Yang et al., 2019). It causes cognitive confusion. When people are confronted with uncertainty stress they tend to be slow to believe new information and adopt preventitive actions, which then makes the problem worse. This study data confirmed that the uncertainty stress from the COVID-19 epidemic has an adverse effect on preventing behaviors needed to reduce the risk of COVID-19 infection.

Controlling uncertainty stress is an important aspect in the prevention of COVID-19 infections. Uncertainty occurs when an event or a situation causes ambiguity, inconsistency, or unpredictability (Mast, 1995). COVID-19 is a new virus, information is unknown or imperfect and there is much ambiguity surrounding the disease. A basic strategy to control uncertain stress is to provide accurate, definitive information in a transparent manner. Definite information comes from established scientific research, but little such research is available with a new and emerging virus like COVID-19. Good research designs and data collection strategies may be difficult to accomplish in middle and low income countries. The best that can happen is to share research findings as quickly as possible with the public keeping in mind that many findings may be preliminary.

All policies and interventions need to be based on scientific evidence when dealing with the COVID-19 epidemic. However, in many areas or regions, policies and interventions were not based on scientific evidence. Many of these policies and interventions were arbitrary and ineffective. The effectiveness of such measures is worth consideration (Scholz, 1983). Ineffective policies and interventions cause people to question their government which may ultimately be detrimental to disease control.

People are in fear and need expert help during the COVID-19 epidemic. However, in order to attract public attention, mass media often invited “famous people” for their opinions even though these people are not well known in area related to the topic included and their thoughts and feelings are not based on science. This can lead to additional uncertainty stress that may hinder prevention efforts among the public. Every expert offering advice and information to the public should be certain, that the information presented is based on scientific evidence. If the media follows this practice it can help to allay fears among the general population and help to reduce the negative impact of the disease.

Ultimately, when communities are faced with uncertainty stress resulting from a disease such as COVID-19 it is natural that they feel fear and despair. The government must then do everything it can to manage the crisis by accurately predicting potential risk, creatively deploying local community resources, and accurately reporting all information to the public.

## Data Availability

Data sharing not applicable to this article because the datasets we used belongs to our Centre for Tobacco Control Research Zhejiang University School of Medicine. Please contact corresponding author for data requests.

